# Large university with high COVID-19 incidence did not increase risk to non-student population

**DOI:** 10.1101/2021.04.27.21255023

**Authors:** Nita Bharti, Brian Lambert, Cara Exten, Christina Faust, Matt Ferrari, Anthony Robinson

**Affiliations:** Center for Infectious Disease Dynamics, Department of Biology, Eberly College of Science, Pennsylvania State University, University Park, PA, USA; College of Nursing, Pennsylvania State University, University Park, PA, USA; GeoVISTA Center, Department of Geography, College of Earth and Mineral Sciences, Pennsylvania State University, University Park, PA, USA

**Keywords:** COVID-19, Universities, colleges, students, pandemic, incidence, public health

## Abstract

Large US colleges and universities that re-opened campuses in the fall of 2020 and the spring of 2021 experienced high per capita rates of COVID-19. Returns to campus were controversial because they posed a risk to the surrounding communities. A large university in Pennsylvania that returned to in-person instruction in the fall of 2020 and spring of 2021 reported high incidence of COVID-19 among students. However, the co-located non-student resident population in the county experienced fewer COVID-19 cases per capita than reported in neighboring counties. Activity patterns from mobile devices indicate that the non-student resident population near the university restricted their movements during the pandemic more than residents of neighboring counties. Preventing cases in student and non-student populations requires different, specifically targeted strategies.

## Introduction

Following the WHO’s March 2020 official declaration of the COVID-19 pandemic, caused by the novel SARS-CoV-2 virus (1), most schools and colleges in the US quickly moved to virtual instruction. To return to campus operations in the fall of 2020, some universities developed COVID-19 policies including frequent testing, mask mandates, distancing in classrooms, and hybrid online and in-person instruction plans (2)(3). Other universities pivoted to fully remote learning and did not return to campus (4). The decision to re-open campuses was controversial because high density student populations would create high-risk transmission situations for students who returned to campus and have negative impacts on their health. Further, outbreaks in students populations had the potential to increase incidence in surrounding communities (5)(6). While individual students could make informed decisions about whether to return to campus based on their risk tolerance, town residents were forced to accept a changing level of local risk based on the decisions of others.

In Centre County, Pennsylvania, The Pennsylvania State University’s (PSU) University Park (UP) campus returned to campus operations in August 2020. The university offered students options for virtual, hybrid learning, and in-person classes with physical distancing in classrooms. Unofficially, the university estimates that about 30,000 undergraduate students returned to the county for the fall semester of 2020, down from approximately 40,000 in the fall of 2019 due to the pandemic. The return to campus quickly resulted in a large COVID-19 outbreak among students (7). We assessed the COVID-19 outbreak in non-student residents in Centre County for comparison to the outbreaks in the surrounding counties, where large universities are not present.

We show that that the timing of the increase in cases among non-student residents of Centre County during the Fall 2020 term lagged the increase in the student population and that the per capita cases among non-student residents of Centre County was lower than in surrounding counties. We then compare activity patterns derived from mobile devices for census block groups within Centre County where students make up more than half the residents, to non-student dominated census block groups in Centre County, and to the neighboring counties.

## Methods

We used publicly available, daily, county-level COVID-19 cases and deaths from the Pennsylvania Department of Health (PA DOH) (https://www.health.pa.gov/topics/disease/coronavirus/pages/cases.aspx)(8)(9) for Centre County and the six neighboring counties with which it shares borders: Blair, Clearfield, Clinton, Huntingdon, Mifflin, and Union (Table 1, Figure 1). Official COVID-19 reporting for these counties began on March 1, 2020 and is ongoing.

**Table 1:**
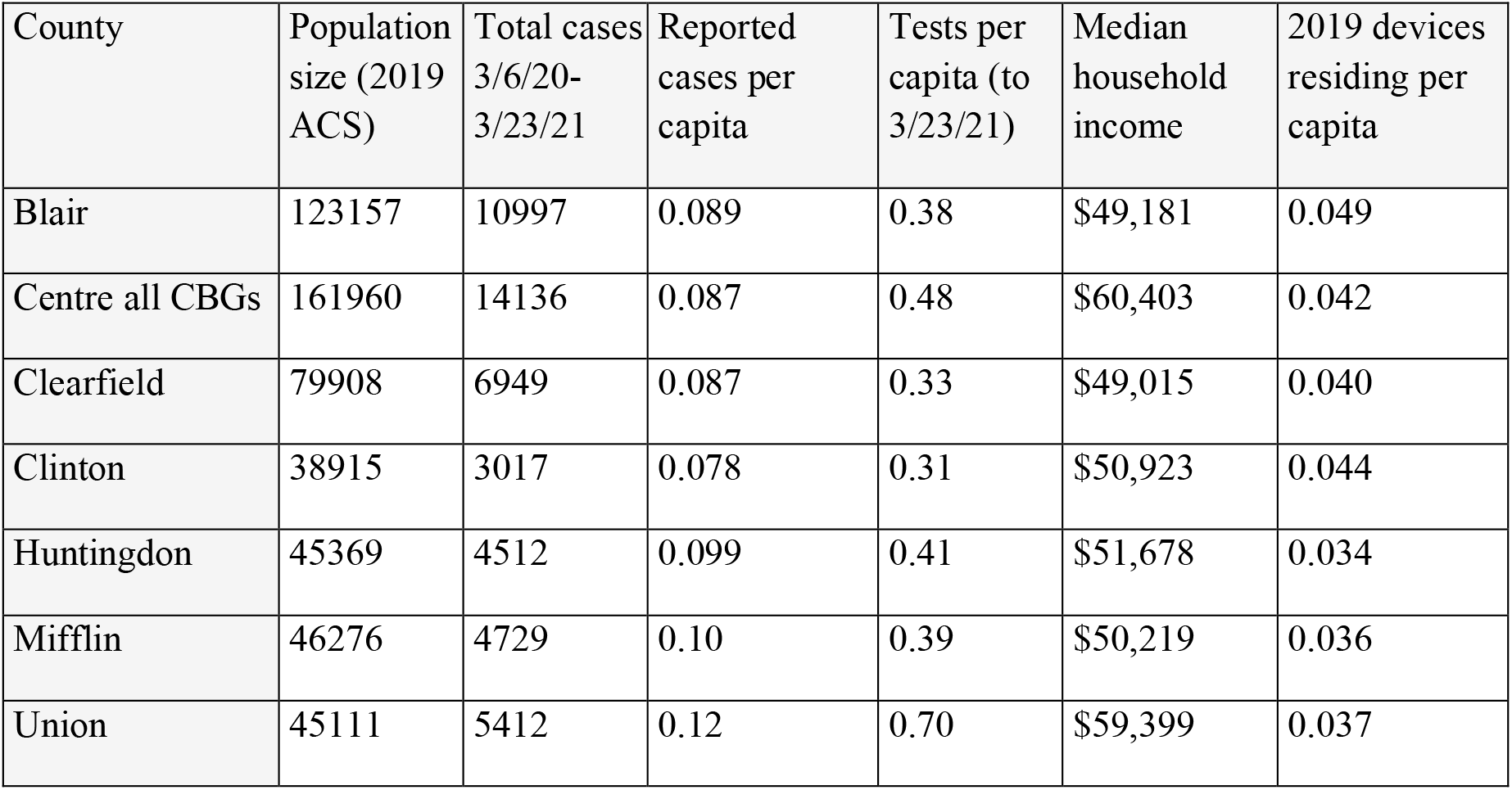
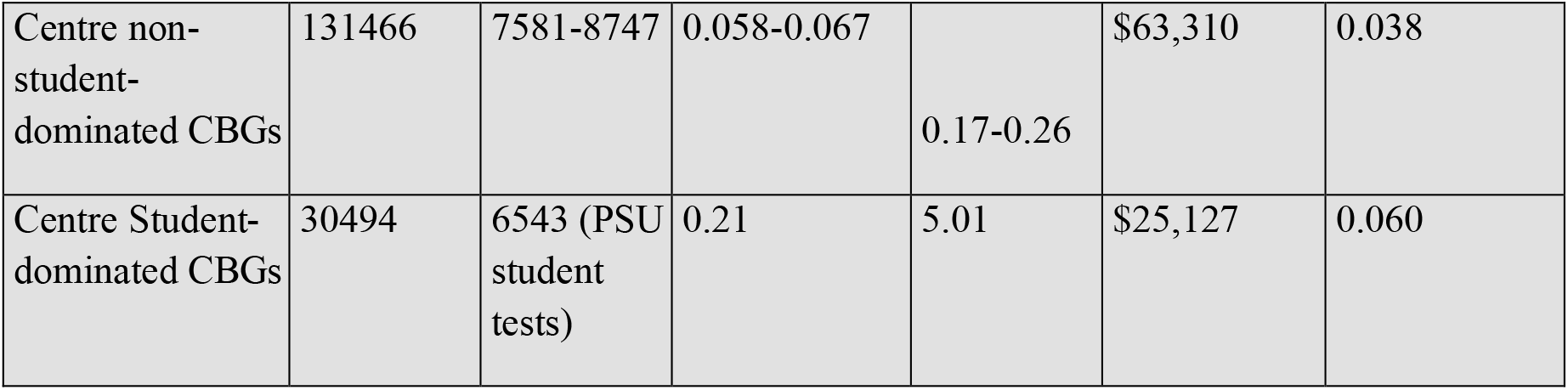
Summary statistics. COVID-19 reporting, census data, SafeGraph mobile-device derived data. *Negative tests for non-students in Centre County cannot be calculated so the number of tests conducted per capita for non-students is not known.

| County | Population size (2019 ACS) | Total cases 3/6/20-3/23/21 | Reported cases per capita | Tests per capita (to 3/23/21) | Median household income | 2019 devices residing per capita |
| --- | --- | --- | --- | --- | --- | --- |
| Blair | 123157 | 10997 | 0.089 | 0.38 | \$49,181 | 0.049 |
| Centre all CBGs | 161960 | 14136 | 0.087 | 0.48 | \$60,403 | 0.042 |
| Clearfield | 79908 | 6949 | 0.087 | 0.33 | \$49,015 | 0.040 |
| Clinton | 38915 | 3017 | 0.078 | 0.31 | \$50,923 | 0.044 |
| Huntingdon | 45369 | 4512 | 0.099 | 0.41 | \$51,678 | 0.034 |
| Mifflin | 46276 | 4729 | 0.10 | 0.39 | \$50,219 | 0.036 |
| Union | 45111 | 5412 | 0.12 | 0.70 | \$59,399 | 0.037 |
| Centre non-student-dominated CBGs | 131466 | 7581-8747 | 0.058-0.067 | 0.17-0.26 | \$63,310 | 0.038 |
| Centre Student-dominated CBGs | 30494 | 6543 (PSU student tests) | 0.21 | 5.01 | \$25,127 | 0.060 |

**Figure 1:**
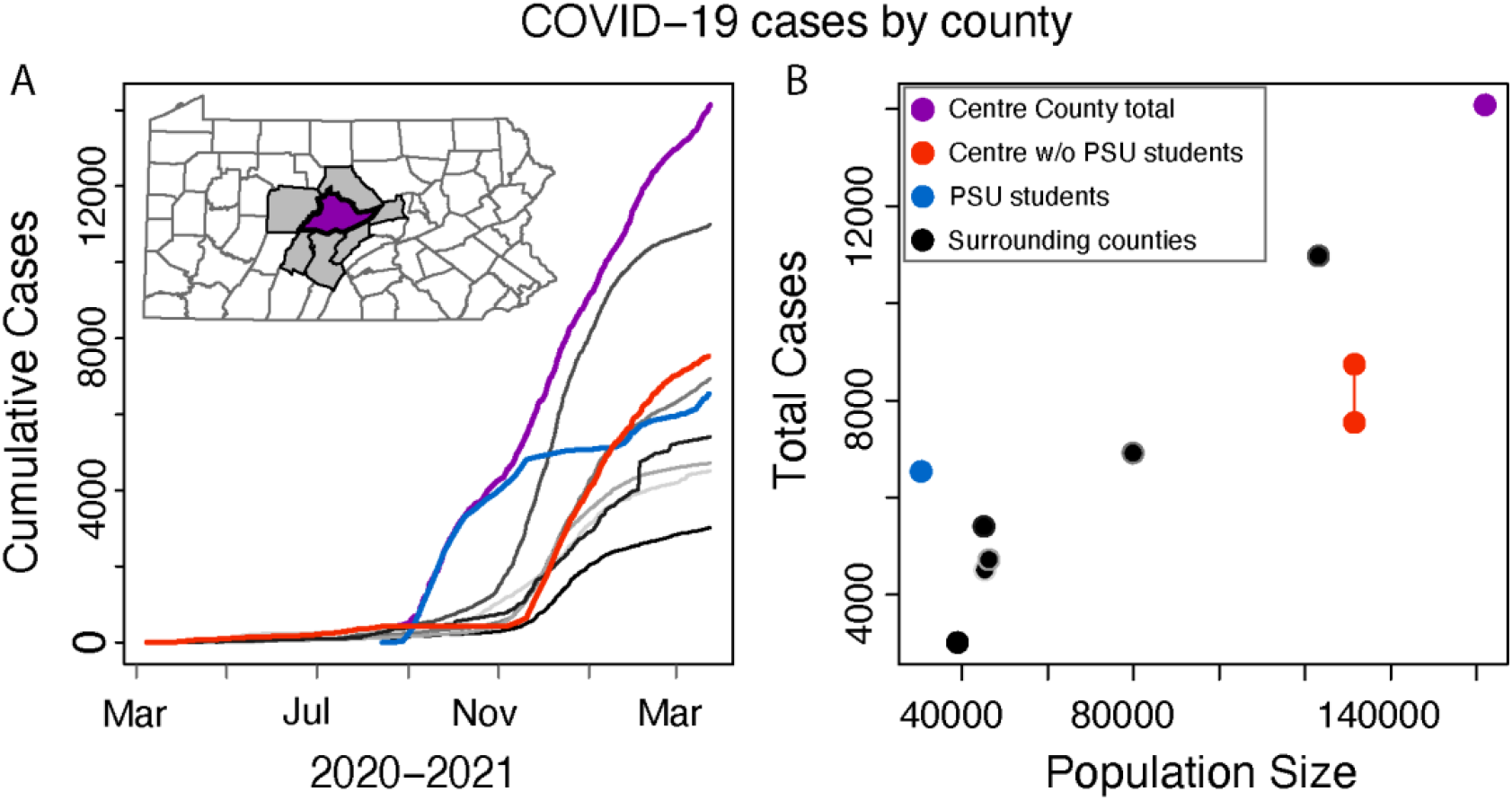
**A**: The cumulative COVID-case trajectory for Centre County minus the student cases (red line) has the same shape as the outbreak for the neighboring counties. When we look at student cases only (blue line), the curve leads other counties. Centre County cumulative cases including the university (purple line) take on the shape of an early increase because of the student cases. **B**: Centre County including PSU student cases (purple dot) reported about the number of cases expected for its population size based on the neighboring counties (black dots). When we separate the student cases the university reported from the non-student residents of the county, Centre County (red dots show possible range of total cases) falls below the number of cases we would expect for the population size. Student cases only (blue dot) are high for the student population size.

Within Centre County, PSU provided COVID-19 testing for UP students from fall 2020 onward and reported anonymized weekly (2020) and daily (2021) confirmed cases, negative test results, and total tests completed for each campus in a public dashboard (https://virusinfo.psu.edu/covid-19-dashboard/)(7). Reporting began on August 7, 2020 and is ongoing.

At the county level, PA DOH reports the total positive, probable, and negative tests for each county. Because PSU is within Centre County, we estimated the number of total positive and negative tests for non-student Centre County residents by subtracting the PSU estimates (from PSU dashboard) from the Centre County estimates provided by PA DOH. However, not all student tests were reported to DOH. A portion of the tests conducted for PSU UP students were completed by a third party vendor, which required student registration. At the time of student registration, an estimated 0-25% of students registered with an address for a family home that did not reflect their residence in Centre County. Their test results were reported to the county of their registered address. This impacts a maximum of 1,166 positive student test results and 10,760 negative student tests. As a result, our estimates of cases and per capita testing among non-student residents in Centre County are imprecise (Table 1).

We also used publicly available data from PA DOH data and PSU to calculate COVID-19 deaths per 100,000 for Centre County, the six neighboring counties, and PSU UP.

We acquired county-level data on median household income, population size, and college enrollment status from the 2019 United Census Bureau’s American Community Survey (ACS) 5-year data (https://www.census.gov/data/developers/data-sets/acs-5year.html) for all seven previously mentioned counties in central PA (10).

We divide the census block groups (CBGs) of Centre County into two categories. We first designated ‘student-dominated CBGs’ as CBGs where >50% of ACS responses report enrollment as undergraduate students. We consider data from the 19 student-dominated CBGs in Centre County to be representative of the student population in Centre County. These 19 CBGs are either on or adjacent to PSU’s UP campus and occupy exactly 6 census tracts. Then, the remaining 25 county census tracts were designated as non-student dominated areas.

SafeGraph (11) receives geolocation data from anonymized mobile devices collected from numerous applications. We analyzed SafeGraph’s mobile device-derived daily visit counts to points of interest (POI), which are fixed locations, such as businesses or attractions. SafeGraph data provide daily counts for total numbers of visits by mobile devices while using at least one application that provides geolocation data to SafeGraph. A “visit” indicates that the device entered the building or spatial perimeter designated as a POI. We acquired daily visit counts for POIs in the seven previously mentioned counties in central PA from January 1, 2019 forward (Table 1) and grouped counts into student-dominated CBGs and non-student dominated CBGs within Centre County. From January 1, 2020 forward, we used SafeGraph data on the median daily minutes that devices spent outside of their home in each county and the student- and non-student dominated CBG divisions in Centre County. The “home location” of each device is defined by its location overnight. Finally, we used SafeGraph’s weekly calculated number of devices residing in each county and the CBG divisions of Centre County for 2019 to measure SafeGraph’s data representation across the seven counties and the CBGs of Centre County.

## Results

Seven clustered counties in central Pennsylvania experienced a similar number of COVID-19 cases per capita through March 23, 2021 (Table 1, Figure 1). In Centre County, the epidemic surged in the student population early in the fall, when students returned to campus in August 2020, and lagged in all neighboring counties. Throughout the fall of 2020, the outbreak in non-students in Centre County lagged the student outbreak by approximately 7-8 weeks in cumulative reported cases, despite the non-student population size being over four times greater than the student population (Figure 1A). Per capita case totals in the student population were greater than all neighboring counties, including the non-student residents of Centre County (Figure 1B, Table 1). Per capita COVID-19 cases were lower among the non-student residents of Centre County than in neighboring counties. Median household income was not correlated to per capita COVID-19 cases at the county level (Pearson correlation = 0.41). Through March 23, testing per capita among PSU students was 5.01, higher than any of the seven counties, which reported per capita testing rates ranging from 0.31 to 0.70 (Table 1). Through March 23, 2021, the mean number of deaths per 100,000 across these seven counties was 219.77 (range 131.51, 378.17); Centre County, including students and non-students, reported the lowest value. We subtract the student population and reported student deaths (N=0) from the Centre County totals and calculate that Centre County non-students experienced 162.02 COVID-19 deaths per 100,000.

In 2019, SafeGraph collected data in Centre County on 4,976,706 visit counts to 2,199 points of interest from a weekly mean of 6,798 mobile devices (range 4,780-8,695). SafeGraph’s 2019 mean count of mobile devices residing in each county represents 4% (range 3.4-4.9%) of the 2019 census population of each county. The Pearson correlation between the mean count of devices residing in a county and 2019 ACS population size of the county was 0.99 for the seven counties (Table 1, Figure 2A). Some demographic groups are likely better represented among SafeGraph’s mobile device users than others; Centre County’s student-dominated CBGs had greater SafeGraph representation (6%) than any other areas in this study (Table 1, Figure 2A). SafeGraph representation was not biased by median household income at the county level (Pearson correlation = -0.20).

**Figure 2:**
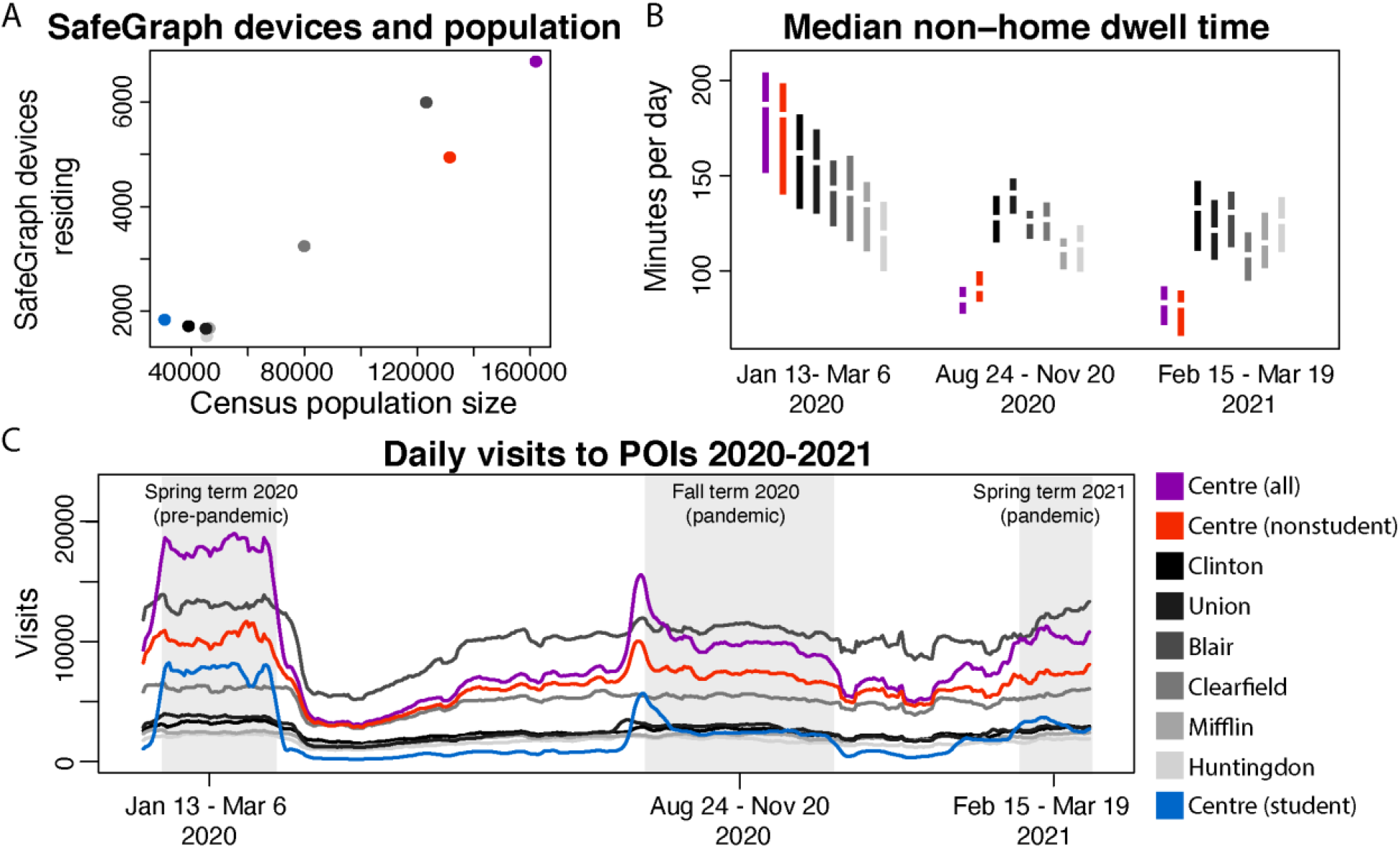
**A**: SafeGraph mean devices per county in 2019 against population size 2019. **B**: Median daily minutes spent outside the home time per county and non-student and student dominated CBGs of Centre County. Mean in center and central 50% in colored bars. From left to right: pre-pandemic, fall term during campus operations, spring term during campus operations. **C**: Rolling seven day mean of total daily visits to points of interest per county and to student-non-student dominated CBGs within Centre County. Areas shaded in grey correspond to time indicated in B. Colors as shown.

In 2019, pre-pandemic SafeGraph data showed different daily visit patterns for the student-dominated CBGs, the rest of Centre County, and the neighboring six counties. The patterns in Centre County strongly reflected academic semesters and university events. The lowest visit counts in student CBGs corresponded to university breaks during winter, summer, and spring while the highest visit counts corresponded to student arrival at the beginning of semesters. This pattern was not seen in the non-student dominated CBGs of Centre County or in other counties (Figure S1). Throughout Centre County, the highest visit counts occurred around home football games, which was not seen in other counties (Figure S1).

SafeGraph data showed reductions in daily visits to POI from March 2020 forward across all seven counties, coinciding with the onset of the pandemic and suggested behavioral restrictions in central PA (12). During the pandemic, small increases in visits were detectable in Centre County just before PSU semesters began in August 2020 for the fall term and February 2021 for the spring term. These increases were driven by the student dominated CBGs of Centre County (Figure 2C). All large university gatherings, intercollegiate athletics, and mid-semester breaks were cancelled. Visit counts across all of Centre County in 2020 were reduced to 56% of their 2019 values and in the non-student dominated CBGs, 2020 visit counts were 71% of their 2019 values. In the student-dominated CBGs, 2020 visit counts were 35% of their 2019 values. Unofficial estimates suspected the undergraduate population returning to Centre County for fall 2020 was 75% of its 2019 level, so the smaller population size alone is unlikely to account for the decline in visit counts. In the neighboring six counties, 2020 visit counts ranged from 81% to 100% of the 2019 values during the fall term period.

During the 2020 spring academic semester from Jan 13 to March 6 2020, before the pandemic restricted movements and impacted behavior in central Pennsylvania, SafeGraph data show mobile devices in Centre County spent more minutes outside the home than devices in any neighboring county (Figure 2B). When the fall semester was in session from August 24 to November 20, 2020 following widespread local pandemic restrictions, the surrounding six counties showed a mean reduction of 3.6-13% in minutes spent outside the home. For the same time period, Centre County showed a mean reduction of 43% in non-student CBGs and 49% in all Centre CBGs in minutes spent outside the home. Mobile devices in Centre County recorded fewer minutes outside the home than any of the surrounding six counties during the fall of 2020. Median household income across the seven counties was correlated to the reduction in minutes spent outside the home between Jan 13 to March 6 2020 and August 24 to November 20, 2020 (Pearson correlation = 0.69) but this relationship was weak when Centre County was removed from the analysis (Pearson correlation = 0.15).

Comparing the fall term of 2020 to the spring term return to campus from February 15 to March 19, 2021, Centre County showed a further 16% reduction in minutes spent outside the home for non-student CBGs and 8% reduction across all Centre CBGs. Comparing across the same time periods, surrounding counties showed between a 12% decrease and 13% increase in minutes spent outside the home.

## Discussion

From March 1, 2020 through March 23, 2021, reported county-level COVID-19 cases per capita in Centre County and six neighboring counties were similar, despite high per capita cases in PSU students (1.8-2.8 times the rate in the 6 surrounding counties). Nearly half of the reported cases from Centre County occurred in university students. The outbreak in students was asynchronous with the outbreak in non-students in Centre County and led the reported cases in the non-student residents of Centre County. The outbreak in Centre County’s non-students was synchronous with outbreaks in surrounding counties. After subtracting student cases reported by PSU, Centre County reported fewer cases per capita than all neighboring counties.

Testing per capita was highest for university students, which may have contributed to higher case detection in students. PA DOH reports showed testing totals were also high for all of Centre County compared to neighboring counties. While we can’t precisely calculate the testing per capita for non-students in Centre County, the number of COVID-19 deaths per capita would not be affected by testing rates. Centre County non-students experienced 162.02 COVID-19 deaths per 100,000, which falls below the mean of 219.77 for these seven counties. This supports the assertion that Centre County nonstudent residents also experienced fewer cases per capita than the surrounding counties and the low number of cases per capita was not an artifact caused by low testing rates.

Mobile-device derived data from SafeGraph showed widespread declines in the number of visits to points of interest (POI) from 2019 to 2020. Centre County recorded greater declines in visit counts from 2019 to 2020 than the surrounding counties. During the pandemic, individuals residing in Centre County spent fewer minutes per day outside their homes during university academic terms than they did for the same time period prior to the pandemic. The decline in minutes spent outside the home in Centre County was greater than the decline in the neighboring counties during the pandemic.

Despite high incidence of COVID-19 among students in Centre County, the delayed outbreak progression in non-student residents of Centre County is consistent with the outbreak timing in neighboring counties, which do not contain large universities. Self-imposed stay-at-home behaviors among Centre County residents may have limited transmission between students and non-student residents of Centre County and slowed transmission among non-student residents (13). Slightly higher median household incomes may have contributed to Centre County residents’ abilities to stay at home during the pandemic more than the residents of neighboring counties (14). Centre County residents may have also disproportionately adopted suggested behavioral interventions, including staying home, in reaction to the large outbreak in Centre County in university students. Ultimately, PSU’s return to campus operations did not cause excess COVID-19 cases per capita in non-student residents around the university but did create a high risk of infection for students who returned to campus.

The primary response to the pandemic through most of 2020 relied on behavioral interventions to prevent SARS-CoV-2 transmission. Public health recommendations emphasized the protective effects of mask wearing, hand washing, and staying home. We show that on average, Centre County residents spent less time outside the home during the pandemic than the residents of the surrounding six counties. Non-student Centre County residents also reported a higher median household income compared to neighboring counties and students in Centre County, which may indicate they were more able to quarantine and work from home. Lower wages among students and in neighboring counties may correspond with higher density housing for students (15) or jobs that cannot transition to remote work in neighboring counties (16). These factors could contribute to increased exposure to SARS-CoV-2 due to less time spent at home (17). County level outbreak prevention and management should consider distinct strategies to meet the specific needs of sub-populations.

## Data Availability

The datasets used that were freely available in the public domain can be found at the links below:
https://www.health.pa.gov/topics/disease/coronavirus/pages/cases.aspx
https://virusinfo.psu.edu/covid-19-dashboard/
https://www.census.gov/data/developers/data-sets/acs-5year.html
Academic researchers must register to receive access to SafeGraph data at no charge for non-commercial purposes only here:
https://www.safegraph.com/academics

## Acknowledgments

We acknowledge SafeGraph for access to their mobile device data, the Pennsylvania Department of Health for making COVID-19 data publicly available at the county level, and The Pennsylvania State University for making their COVID-19 data publicly available. We acknowledge financial support from Penn State University’s Huck Institutes of the Life Sciences and the Institute for Computational and Data Sciences. The funding sources had no role in the study design, execution or interpretation of the results.

## Data availability and ethics Statement

The datasets used that were freely available in the public domain can be found at the links below: https://www.health.pa.gov/topics/disease/coronavirus/pages/cases.aspx; https://virusinfo.psu.edu/covid-19-dashboard/; https://www.census.gov/data/developers/data-sets/acs-5year.html. Academic researchers must register to receive access to SafeGraph data at no charge for non-commercial purposes only here: https://www.safegraph.com/academics. SafeGraph data are anonymized and aggregated.

---

**Figure S1:**
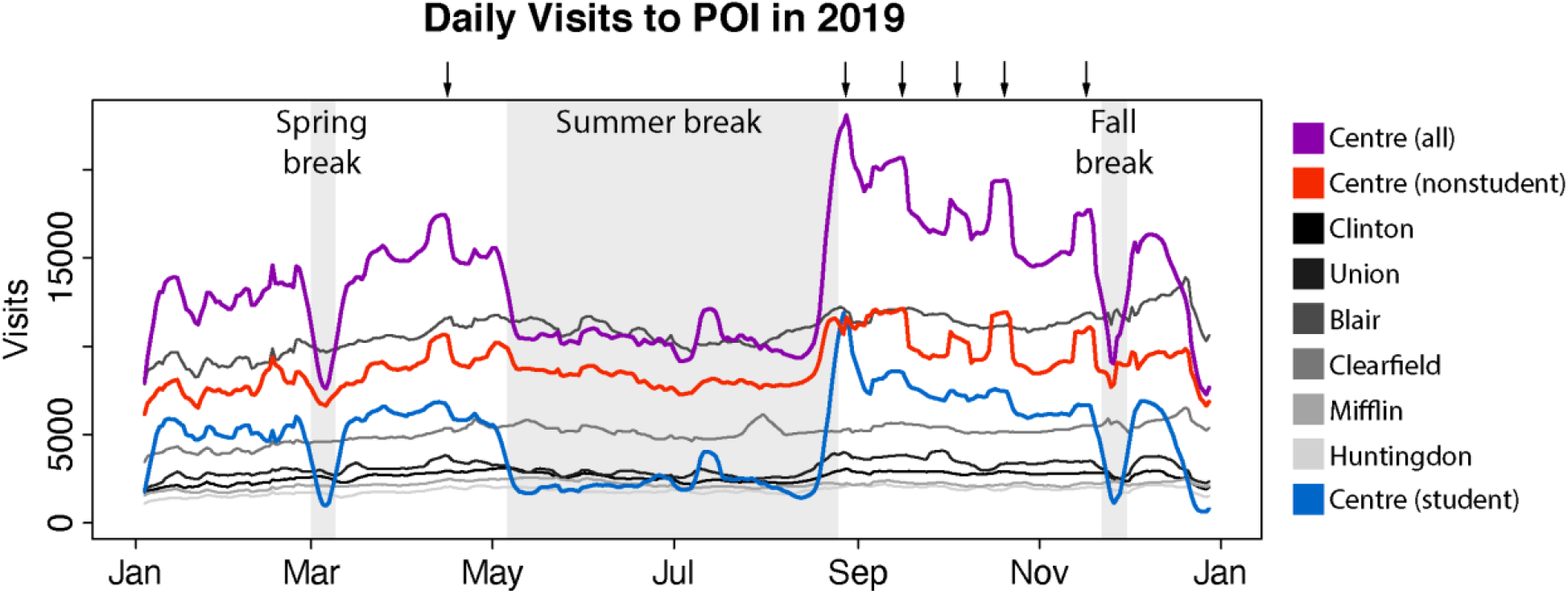
Rolling seven day mean of total daily visits to points of interest per county and student- and non-student dominated CBGs within Centre County. Areas shaded in grey correspond to university breaks, arrows correspond to football games and a ticketed spring scrimmage.

